# Multisystem Symptom Burden and Functional Impairment in a Specialty Benzodiazepine Taper Cohort

**DOI:** 10.1101/2025.10.07.25336923

**Authors:** Valsa S. Madhava

## Abstract

**Background:** Benzodiazepine withdrawal can be prolonged and disabling, with symptoms extending beyond anxiety and insomnia. We characterized symptom burden and functional impairment in a specialty taper cohort and evaluated an exploratory rule for organizing complex, multisystem presentations.

**Methods:** We analyzed cross-sectional intake data from 39 consecutive patients: 37 seeking to begin a supervised taper and two who had completed tapering but remained symptomatic. All 39 returned a 233-item symptom questionnaire, and 38 completed the PROMIS-29 Profile v2.0. Questionnaire items were assigned in advance to five domains: CRH/Adrenergic, Excitatory/Neuroinflammatory, Autonomic, Basal Ganglia–Cerebellar, and MCAS-overlap. Severe symptoms (≥7/10) were counted across the first four domains. Patients with three or fewer severe symptoms were classified as Low-symptom; otherwise, the domain with the highest count determined the dominant-axis label, with ties classified as Mixed. MCAS-overlap was coded separately. The categorization was repeated at severity thresholds from ≥5/10 to ≥8/10.

**Results:** Patients endorsed a median of 74 symptoms (IQR, 48–124), with a median of 24 rated severe (IQR, 7–42). Thirty-five of 39 patients (89.7%) had severe symptoms in at least two core domains. PROMIS-29 mean T-scores were elevated for Anxiety, Depression, Sleep Disturbance, and Fatigue and were lower for Physical Function and Ability to Participate in Social Roles and Activities; Pain Interference was close to the population mean. At the primary ≥7/10 severity threshold, 14 patients were classified as CRH/Adrenergic, 14 as Excitatory/Neuroinflammatory, four as Autonomic, and seven as Low-symptom; none were classified as Basal Ganglia–Cerebellar or Mixed. Twenty-two patients (56.4%) met the operational criterion for MCAS-overlap. Agreement with the primary labels was 87.2% at ≥6/10 and 69.2% at both ≥5/10 and ≥8/10 severity thresholds.

**Conclusions:** This cohort had severe, multisystem symptom burden and substantial functional impairment. The five-domain framework provided a structured way to organize these presentations, but the dominant-axis labels were threshold-dependent and should be interpreted as exploratory summaries rather than discrete patient subtypes. Larger longitudinal studies using validated instruments and data-driven methods are needed to determine whether stable symptom subgroups exist.

**Version Note:** Version 4 revises the interpretation of the rule-based symptom categories and corrects several descriptive results following re-review of source records, including PROMIS-29 scoring, demographics, exposure, and category counts. The cohort and underlying source data are unchanged. The corrected results continue to show severe, multisystem symptom burden and substantial functional impairment, and the categories are now reported as exploratory clinical summaries rather than discrete patient subtypes.

## Introduction

Benzodiazepines remain widely prescribed, yet long-term use and discontinuation can present substantial clinical challenges. Withdrawal may be prolonged, severe, and clinically heterogeneous, with psychiatric, neurological, autonomic, gastrointestinal, sensory, cognitive, affective, and immune-related symptoms. This breadth and variability have been documented in large patient surveys of benzodiazepine withdrawal and enduring symptoms [1,2].

Benzodiazepine-induced neurological dysfunction (BIND) has been proposed as a descriptive construct for symptoms and associated functional consequences that may emerge during benzodiazepine use or tapering and continue after discontinuation [3]. BIND emphasizes that benzodiazepine-related symptoms may extend beyond short-term rebound anxiety or insomnia, but it does not explain why presentations may be prolonged, multisystem, or disabling. Research on GABAₐ receptor adaptations, tolerance, and circuit-level changes provides a neurobiological context for withdrawal after long-term benzodiazepine exposure [4–6]. However, less is known about how complex symptom presentations can be organized clinically.

The specialty taper program described here used structured intake assessments to document symptoms and functioning. A 233-item questionnaire recorded the presence and severity of a broad range of symptoms, while the Patient-Reported Outcomes Measurement Information System 29-item Profile (PROMIS-29) v2.0 provided a standardized, population-referenced assessment of symptom and functional domains [7].

To organize the questionnaire, symptoms were assigned to four core domains informed by physiological and clinical literature: CRH/Adrenergic, Excitatory/Neuroinflammatory, Autonomic, and Basal Ganglia–Cerebellar. A fifth domain, MCAS-overlap, was treated as a cross-cutting modifier rather than a competing axis. These domains were defined before the dominant-axis rule was applied to individual patients.

Using intake data from 39 consecutive patients, this study aimed to describe overall symptom burden and functional impairment, characterize the distribution of severe symptoms within the five-domain framework, and evaluate the sensitivity of the resulting dominant-axis labels to the severity threshold applied.

## Methods

### Clinical Setting and Cohort

This study analyzed observational data from 39 consecutive patients enrolled in a specialty benzodiazepine taper program. All patients enrolled during this period were included; no selection was made on the basis of symptom pattern or severity. Thirty-seven presented with the intention to begin a supervised taper, and two had already completed benzodiazepine tapering but remained symptomatic at enrollment.

The present analysis used intake data from the 233-item symptom questionnaire and the PROMIS-29. Patients completed these assessments at baseline, before this program provided any treatment. Standard medical and substance-use histories were collected during the same structured intake process. Other baseline forms, including the diet diary, nutrient-deficiency questionnaire, and ACE questionnaire, informed clinical care only and did not contribute to the analyses reported here.

### Data Collection and Instruments

#### Symptom Questionnaire Construction and Provenance

A 233-item symptom questionnaire was constructed from the Benzodiazepine Information Coalition’s “Withdrawal & Post-Withdrawal Symptoms” list [8], with symptom wording largely preserved. Wording was lightly standardized, such as harmonizing dashes and writing out “and.” These formatting changes did not affect axis item counts or dominant-axis labels. All patients returned the questionnaire at intake. It was in use from the program’s inception, so all patients returned the same baseline assessment.

As part of the present analysis, we retrospectively reconciled the administered questionnaire with the source list. Of the 243 source symptom phrases, 239 were represented in the questionnaire; four—burping, catatonia, muscle rigidity, and muscle stiffness—had been inadvertently omitted. Seven administered items combined two source phrases each, resulting in 232 unique item labels. Because one item was inadvertently duplicated, the administered questionnaire contained 233 rated rows. These discrepancies were transcription artifacts, not planned modifications. The full derivation is shown in Appendix 1.

#### Rating Scale

Each item was rated on a 0–10 severity scale (0 = not at all; 10 = most severe). Scores ≥7/10 were used as the analytic threshold for a “severe” symptom. This threshold reflected a clinical convention used within the program to identify the subset of a 233-item instrument most likely to warrant clinical attention.

The threshold is an operational convention rather than a psychometrically derived value; no external standard establishes ≥7/10 as a validated boundary for clinical significance. Because the rule-based categorization described below depends on it, its influence is examined directly in a threshold-sensitivity analysis. Throughout this manuscript, we refer to symptoms rated ≥7/10 as “severe” for simplicity.

For the Mast Cell Activation Syndrome (MCAS)-overlap modifier, a lower severity threshold of ≥5/10 was used. This was intended to capture features overlapping with those reported in MCAS, rather than to establish a formal MCAS diagnosis.

#### PROMIS-29

The PROMIS-29 v2.0 questionnaire was completed at intake by 38 of the 39 patients. It assessed seven domains — Physical Function, Anxiety, Depression, Fatigue, Sleep Disturbance, Ability to Participate in Social Roles and Activities, and Pain Interference — plus a single Pain Intensity item rated from 0 to 10. Each domain included four Likert-scale items. Raw scores were converted to T-scores (mean = 50, SD = 10) using the official PROMIS-29 v2.0 short-form conversion tables [7]. Higher scores reflect greater symptom burden in Anxiety, Depression, Sleep Disturbance, Fatigue, and Pain Interference, and better functioning in Physical Function and Ability to Participate in Social Roles and Activities.

PROMIS-29 domain scores were derived from the source questionnaires. Reverse-coded items— Sleep Disturbance item 18 and all four Ability to Participate items—were scored in the direction specified by the instrument. When a source questionnaire included more than one dated administration, we used the baseline administration. Domains containing unanswered items were not scored for that patient, and domain-specific sample sizes are reported in Table 3. Details of response reconciliation and scoring procedures are provided in Supplement 1.

#### Benzodiazepine and Z-Drug Exposure

Benzodiazepine and Z-drug exposure at intake was ascertained from the clinical record. Where intake medication or dose information was incomplete, exposure was reconstructed from documented sources, including intake forms, medication records, first-visit notes, correspondence, and the state prescription drug monitoring program; the source used was recorded for each patient.

Daily doses of benzodiazepines and Z-drugs — the latter including zolpidem, zopiclone, and zaleplon — were converted to diazepam equivalents using Ashton Manual equivalency estimates, which are approximate rather than exact [9]. For patients taking more than one benzodiazepine, the diazepam-equivalent doses of the individual agents were summed.

#### Medical and Substance Use History

Patients completed structured medical and substance-use history forms at intake. Self-reported health problems and concerns were collected as part of the intake medical history and grouped descriptively by body system (Appendix 3); substance use is summarized in Appendix 4. These entries were used clinically and were not independently verified or adjudicated as diagnoses.

#### Data Completeness

All 39 patients returned the 233-item symptom questionnaire and provided analyzable data; 38 patients completed the PROMIS-29, 38 completed the medical history, and 36 completed the substance use history (Appendix 6).

Unanswered symptom-questionnaire items were treated as zero in the analysis matrix and therefore could not increase any axis count. Patients were retained in the exploratory dominant-axis summary regardless of the proportion of unanswered items.

### Five-Domain Framework

The five domains were derived by reviewing the questionnaire items together with the physiological processes plausibly related to them. They were defined before the rule-based categorization was applied to any patient. The Discussion considers the biological rationale and supporting literature. In this manuscript, “domain” refers to one of the five predefined symptom groupings. “Axis” refers specifically to a domain used as a counting unit in the dominant-axis categorization. The first four domains served as the four competing core axes; MCAS-overlap was treated separately as a cross-cutting modifier. The term “axis” does not denote a demonstrated physiological circuit.

The five domains, with representative symptoms, were:

- **CRH/Adrenergic:** anxiety, insomnia, tachycardia, hypervigilance, sweating.
- **Excitatory/Neuroinflammatory (ENI):** fatigue, brain fog, diffuse pain, sensory hypersensitivity.
- **Autonomic:** orthostatic intolerance, palpitations, variable blood pressure or heart rate.
- **Basal Ganglia–Cerebellar (BG/Cer):** tremor, muscle spasm, coordination difficulty.
- **MCAS-overlap:** flushing, gastrointestinal reactivity, histamine-related sensitivities.

Each of the 233 questionnaire items was reviewed individually and assigned to the single domain considered the best fit based on the proposed physiological processes. Appendix 1 provides the complete item-by-domain assignment. The resulting counts reflect the number of questionnaire items assigned to each domain, not an attempt to balance the domains: CRH 47, ENI 85, Autonomic 61, BG/Cer 19, and MCAS 21. The four core axes were compared to assign the dominant-axis label; MCAS-overlap was treated as a cross-cutting modifier and did not compete for that label. The unequal item counts across axes bear on the interpretation of the resulting categories and are examined in the Discussion.

Together, these domains provide an organizing framework for the symptom set. They are not claimed to constitute a complete, minimal, or biologically validated set of mechanisms.

### Rule-Based Dominant-Axis Categorization

Symptoms rated ≥7/10 were operationally classified as severe, reflecting the program’s clinical convention for prioritizing the most intense symptoms. Patients with three or fewer severe symptoms across the four core axes were classified as Low-symptom, regardless of their distribution. For all other patients, severe-symptom counts were compared across the four core axes. The axis with the highest count determined the dominant-axis label, and ties between two or more axes were classified as Mixed.

Features overlapping with those reported in MCAS were coded separately using the MCAS-overlap modifier, defined as two or more of the 21 MCAS-overlap symptoms rated ≥5/10 in severity (see Appendix 2 for symptom tiers). The modifier could co-occur with any dominant-axis label and was not included in the comparison used to assign that label. This operational definition does not imply a formal MCAS diagnosis.

The ≥7/10 severity threshold, the cutoff of three for the Low-symptom category, and the ≥5/10 threshold for MCAS-overlap were investigator-defined operational choices and were not derived from external standards.

This procedure applies a rule to symptom counts. It is not a clustering procedure, latent-class analysis, or other method designed to identify data-driven groups, and no analysis was performed to test whether discrete groups existed.

### Threshold-Sensitivity Analysis

To assess the sensitivity of the dominant-axis labels to the primary ≥7/10 severity threshold, we repeated the full categorization procedure using severity thresholds of ≥5/10, ≥6/10, and ≥8/10. At each tested threshold, we counted symptoms meeting that threshold separately within each of the four core axes. Patients with three or fewer qualifying symptoms across the four core axes were classified as Low-symptom. For all other patients, the axis with the highest count determined the dominant-axis label, and ties were classified as Mixed. Labels obtained at each tested threshold were compared with the primary labels generated at ≥7/10. Agreement was defined as the proportion of the 39 patients who received the same label at the tested threshold and at ≥7/10. The MCAS-overlap modifier was not included in this analysis.

### Statistical Analysis

Analyses were descriptive only. Given the exploratory nature of the study and the modest sample size, no inferential statistical testing was performed.

### Ethics and IRB Approval

The Biomedical Research Alliance of New York (BRANY) determined this study to be exempt from IRB review and granted a full waiver of HIPAA authorization. All analyses were performed using de-identified data.

## Results

### Cohort Characteristics, Medication Exposure, Symptom Burden, and Functional Impairment

#### Demographic Characteristics

Demographic information was available for 38 of 39 patients (Table 1).

**Table 1.** Cohort demographics (N = 38).

| Characteristic | Value |
| --- | --- |
| Age at intake, years | Mean 48.9 (SD 14.5), range 21.9–76.3 |
| Sex | Female 20 (52.6%); Male 18 (47.4%) |
| Race | White 28 (73.7%); Black or African American 1 (2.6%); Asian 1 (2.6%); Asian and White 1 (2.6%); Declined to specify 1 (2.6%); Missing 6 (15.8%) |
| Ethnicity | Not Hispanic or Latino 27 (71.1%); Hispanic or Latino 4 (10.5%); Declined to specify 1 (2.6%); Missing 6 (15.8%) |
| Marital status | Single 17 (44.7%); Married 15 (39.5%); Other 2 (5.3%); Missing 4 (10.5%) |
| Employment status | Employed 19 (50.0%); Retired 7 (18.4%); Unemployed 6 (15.8%); Student 2 (5.3%); Missing 4 (10.5%) |

*Note:* Demographic data were available for 38 of the 39 patients. Percentages are calculated using N = 38; missing and declined responses are shown explicitly rather than excluded from denominators. Characteristics were recorded on the intake health history form and are reported as captured; they were not recoded into binary categories. Age at intake was calculated from date of birth and date of program entry.

Mean age at intake was 48.9 years (SD 14.5, range 21.9–76.3). Twenty patients (52.6%) were female and 18 (47.4%) male. Twenty-eight patients (73.7%) identified as White and 27 (71.1%) as not Hispanic or Latino; race and ethnicity were each not recorded for six patients (15.8%). Seventeen patients (44.7%) were single and 15 (39.5%) married.

Nineteen patients (50.0%) were employed at intake, seven (18.4%) were retired, six (15.8%) unemployed, and two (5.3%) students; employment and marital status were each not recorded for four patients (10.5%).

#### Benzodiazepine and Z-Drug Exposure

Among the 37 patients taking a benzodiazepine or Z-drug at intake, alprazolam was the most common primary agent (11/37, 29.7%), followed by clonazepam (10/37, 27.0%), lorazepam (8/37, 21.6%), diazepam (7/37, 18.9%), and a Z-drug (1/37, 2.7%) (Table 2).

**Table 2.** Benzodiazepine and Z-drug exposure at intake (N = 37).

| Characteristic | Value |
| --- | --- |
| **Primary benzodiazepine or Z-drug, n (%)** |  |
| Alprazolam | 11 (29.7) |
| Clonazepam | 10 (27.0) |
| Lorazepam | 8 (21.6) |
| Diazepam | 7 (18.9) |
| Z-drug | 1 (2.7) |
| **Diazepam-equivalent dose, mg/day** |  |
| Mean (SD) | 27.0 (24.6) |
| Median (IQR) | 18.0 (10.0–40.0) |
| Range | 0.4–100 |
| **Duration of use, years (n = 36)** |  |
| Mean (SD) | 9.0 (8.2) |
| Median (IQR) | 7.0 (2.8–14.0) |
| Range | 0.17–34.0 |

*Note:* Exposure data were available for 37 of 39 patients; the two patients not taking a benzodiazepine or Z-drug at enrollment were excluded. Duration of use was available for 36. Daily doses were converted to diazepam equivalents using the Ashton Manual [9]. Four patients were taking a second agent in addition to their primary agent; for these patients, the equivalents were summed to give a total daily dose. The agent shown above is the primary agent recorded at intake. Z-drugs (zolpidem, zopiclone, zaleplon) are non-benzodiazepine hypnotics; conversions followed Ashton Manual equivalencies.

The mean diazepam-equivalent dose was 27.0 mg/day (SD 24.6; median 18.0, IQR 10.0–40.0; range 0.4–100). Duration of use was available for 36 patients and averaged 9.0 years (SD 8.2; median 7.0, IQR 2.8–14.0; range 0.17–34.0).

#### Medical and Substance-Use History

Among the 38 patients who provided medical histories, all reported at least one psychiatric or emotional concern, a broad self-reported category that included items such as anxiety, depression, history of trauma or abuse, and lack of a support system. Neurological problems were reported by 27/38 (71.1%) and immune/autoimmune-related problems by 22/38 (57.9%); additional reported problems across endocrine/metabolic, gastrointestinal, musculoskeletal, respiratory, and cardiovascular categories are summarized in Appendix 3.

Substance-use histories are summarized in Appendix 4.

#### Overall Symptom Burden

All 39 patients returned the 233-item symptom questionnaire. Across the cohort, responses indicated substantial symptom burden. Patients endorsed a median of 74 symptoms (IQR 48–124; range 14–213) and had a median of 24 symptoms rated severe (≥7/10; IQR 7–42; range 0–136). Fourteen of 39 patients endorsed more than 100 symptoms, and 21 of 39 had at least 20 symptoms rated severe. The symptoms most frequently rated severe were anxiety (27/39), insomnia (24/39), hypersensitivity to stress (21/39), loss of confidence (18/39), nervousness (18/39), and exhaustion (17/39). Severe symptoms were distributed across domains rather than confined to one: 35 of 39 patients (89.7%) had at least one severe symptom in two or more of the four core domains, 29 (74.4%) in three or more, and 23 (59.0%) in all four.

Item-level missingness on the questionnaire was low for most patients (median 1.3%; IQR 0.6%–5.0%), although three patients left more than 60% of items unanswered (62.1%, 64.2%, and 69.0%). Because unanswered items were treated as zero, symptom and axis counts for these patients may be underestimated, and their dominant-axis labels should be interpreted cautiously. This concern was greatest for one patient, whose highest and second-highest axis counts were four and three, respectively. Substantial missingness was not present in the Low-symptom group: each of the seven patients assigned this label left no more than 1.7% of questionnaire items unanswered.

#### PROMIS-29 Symptom and Function Scores

Standardized symptom and functional domains were assessed using the PROMIS-29, an externally normed instrument completed by 38 of the 39 patients at intake.

As shown in Table 3 and Figure 1, mean T-scores were elevated relative to the population-referenced mean of 50 for Anxiety (mean 68.8, SD 7.8), Depression (62.3, SD 9.7), Sleep Disturbance (61.0, SD 8.5), and Fatigue (60.7, SD 10.6). Pain Interference was close to the population mean (51.5, SD 12.4). Mean scores were lower for Physical Function (45.5, SD 10.7) and Ability to Participate in Social Roles and Activities (40.7, SD 7.5), indicating reduced functioning in these domains.

**Figure 1.**
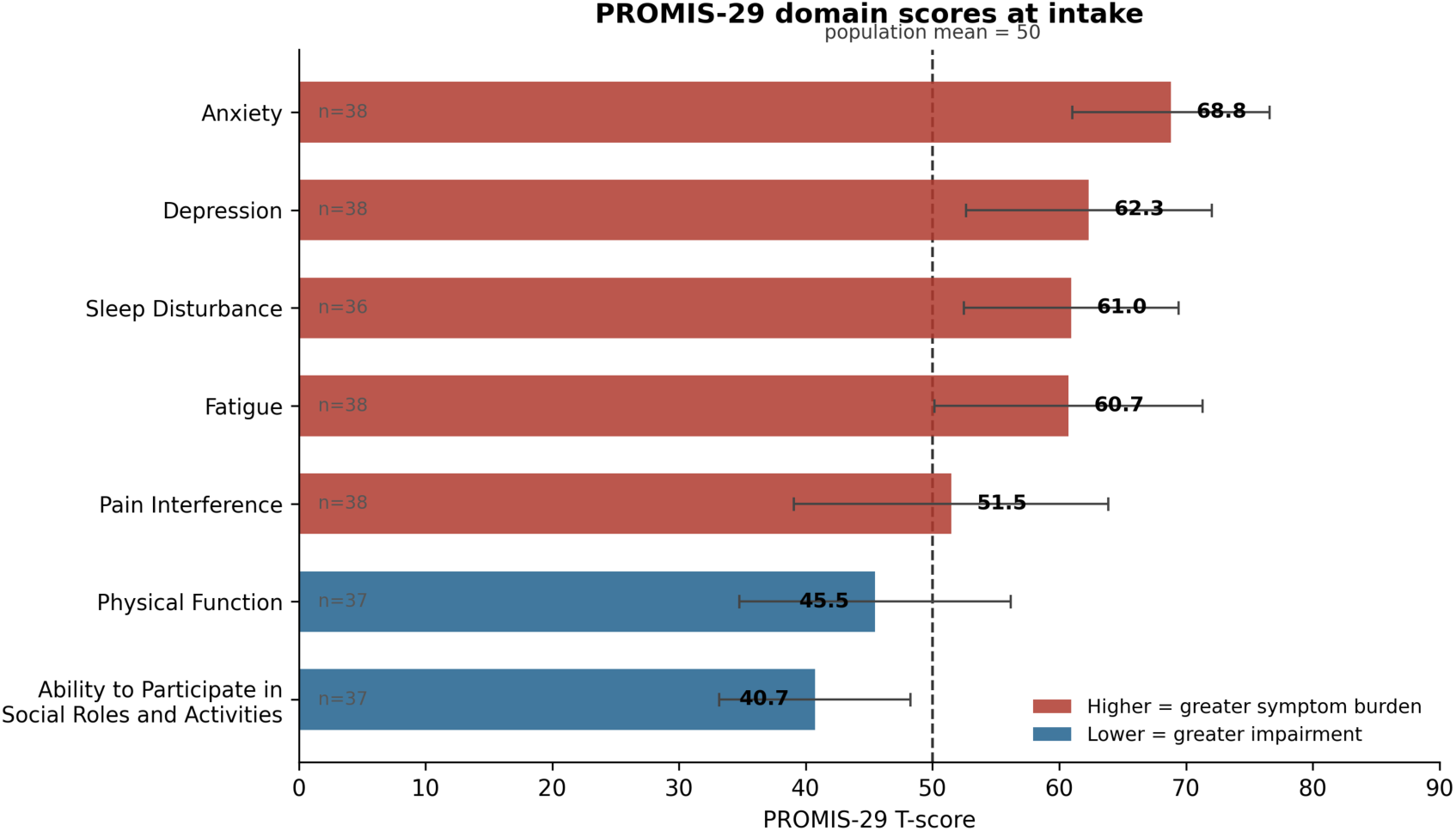
PROMIS-29 domain scores at intake. T-scores are normed to a population mean of 50 and standard deviation of 10. Higher scores indicate greater symptom burden for Anxiety, Depression, Sleep Disturbance, Fatigue and Pain Interference; lower scores indicate greater impairment for Physical Function and Ability to Participate in Social Roles and Activities. Error bars show one standard deviation. Domain-specific sample sizes are given in Table 3.

**Table 3.**
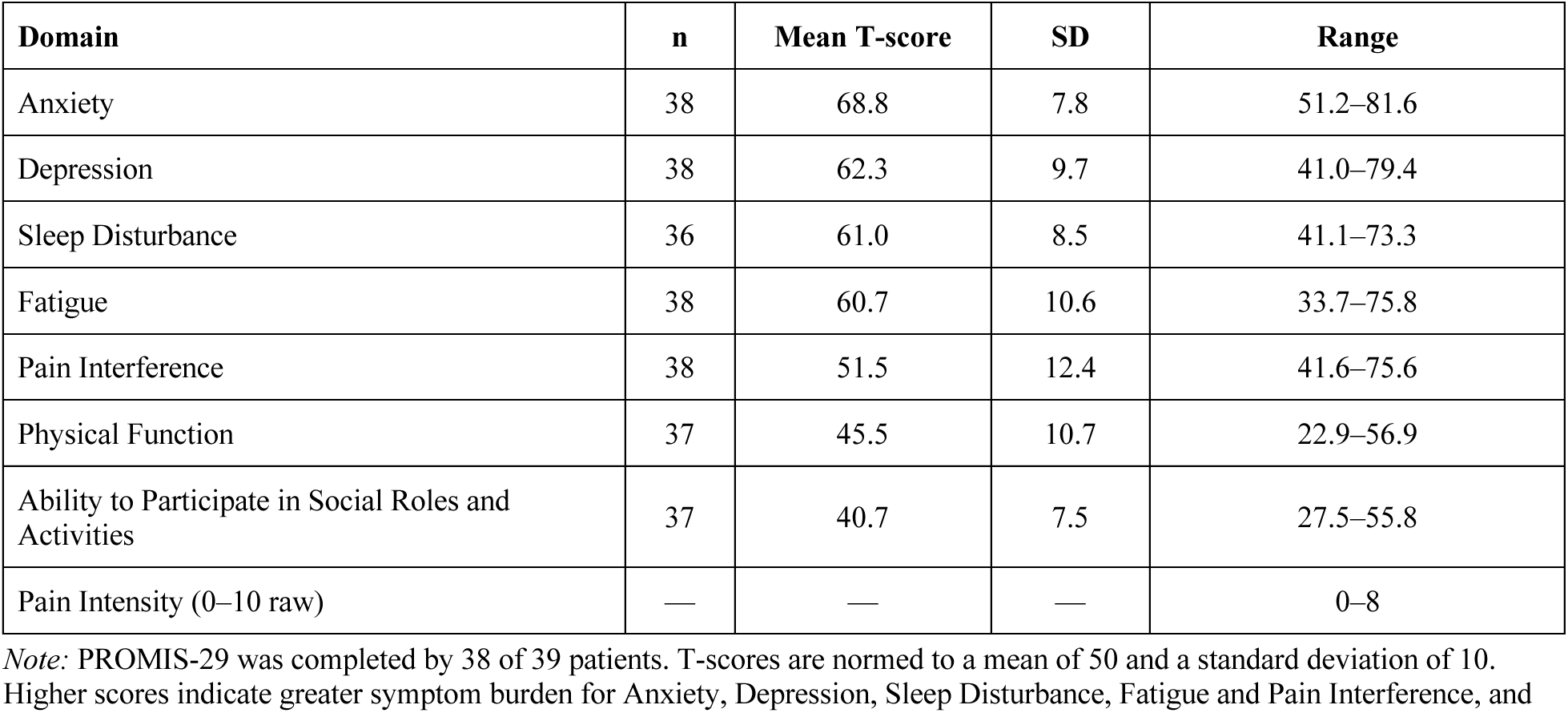
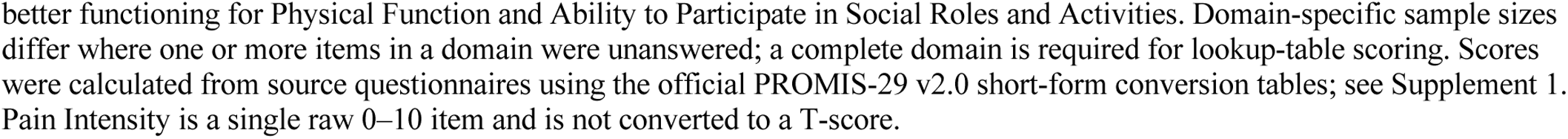
PROMIS-29 v2.0 outcomes at intake.

Reported Pain Intensity scores ranged from 0 to 8 on the 0–10 scale. Domain scores were available for 38 patients except for Sleep Disturbance (n = 36), Physical Function (n = 37), and Ability to Participate in Social Roles and Activities (n = 37), owing to unanswered items.

### Rule-Based Dominant-Axis Categorization

All 39 patients were categorized by counting symptoms rated ≥7/10 severity across the four core axes: those with three or fewer severe symptoms were classified as Low-symptom; otherwise, the axis with the highest count determined the dominant-axis label, with ties classified as Mixed. Full details of the categorization procedure are provided in Methods.

The resulting distribution is shown in Table 4 and Figure 2; patient-level tallies are provided in Appendix 5. CRH/Adrenergic and Excitatory/Neuroinflammatory were the most frequent labels, each assigned to 14 patients (35.9%). Four patients received the Autonomic label (10.3%), and none received the Basal Ganglia–Cerebellar label. Seven patients were classified as Low-symptom (17.9%), and none met the Mixed criterion at the ≥7/10 severity threshold.

**Figure 2.**
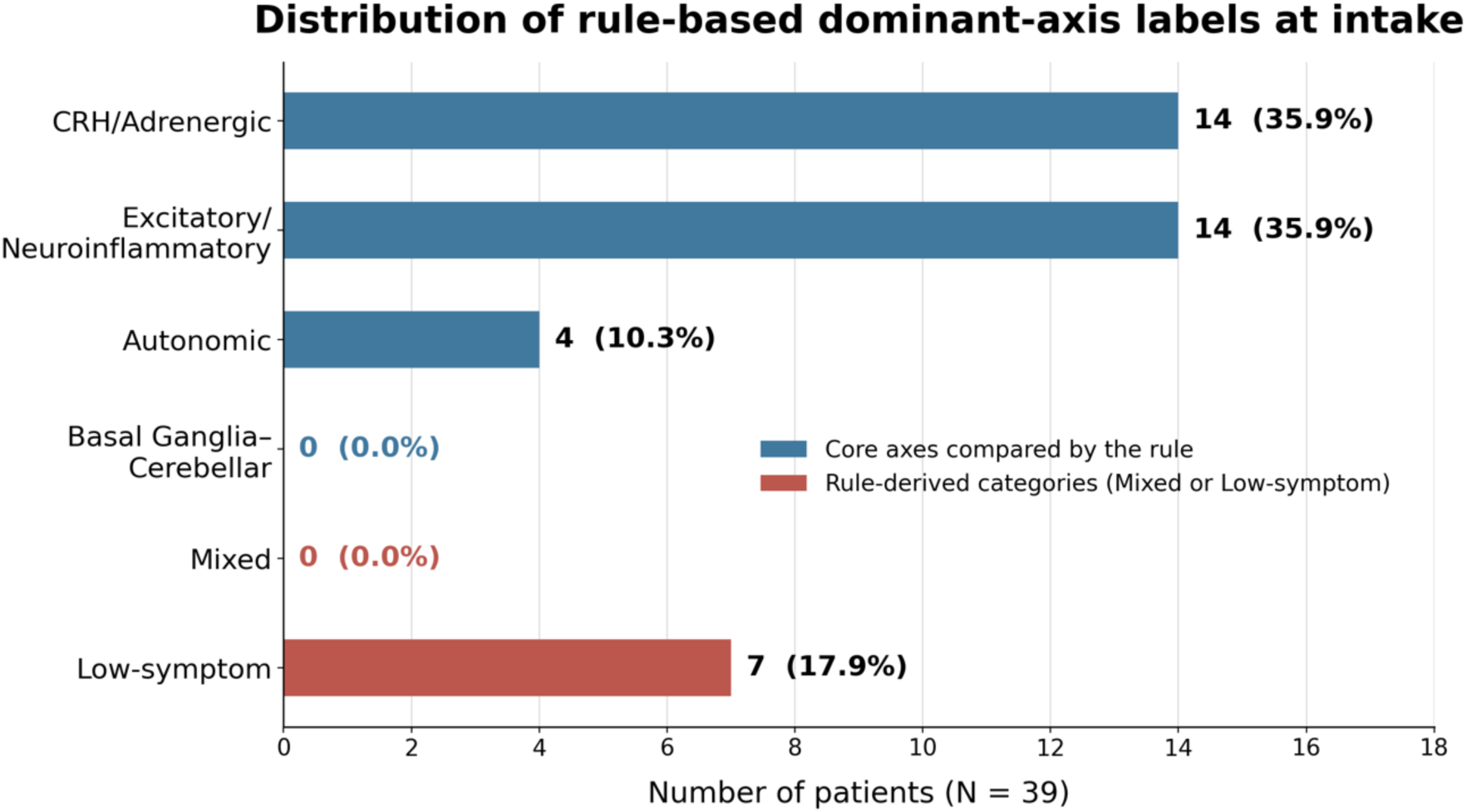
Distribution of rule-based dominant-axis labels at intake (N = 39). The labels are exploratory summaries generated by the rule and do not establish discrete patient subtypes. For patients not classified as Low-symptom, the label identifies the predefined core axis containing the highest number of symptoms rated ≥7/10; ties were classified as Mixed. Sensitivity to the severity threshold is reported in Table 5, and the implications of unequal axis item counts are considered in the Discussion.

**Table 4.**
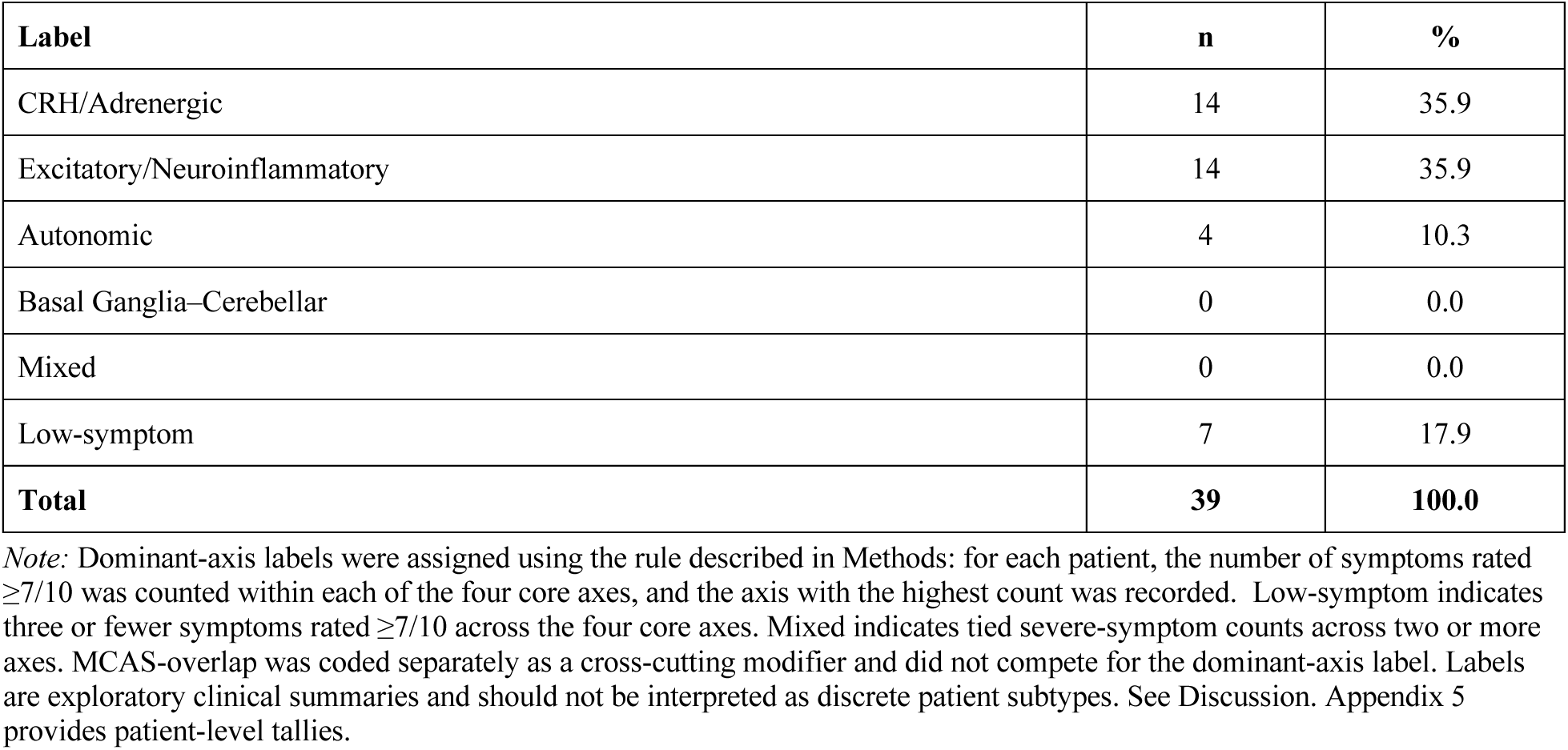
Rule-based dominant-axis labels at intake (N = 39).

**Table 5.** Dominant-axis labels by severity threshold (N = 39).

| Label | ≥5 | ≥6 | ≥8 |
| --- | --- | --- | --- |
| CRH/Adrenergic | 15 | 13 | 19 |
| Excitatory/Neuroinflammatory | 17 | 16 | 6 |
| Autonomic | 2 | 2 | 2 |
| Basal Ganglia–Cerebellar | 0 | 0 | 0 |
| Mixed | 3 | 2 | 1 |
| Low-symptom | 2 | 6 | 11 |
| **Agreement with ≥7/10** | **27/39 (69.2%)** | **34/39 (87.2%)** | **27/39 (69.2%)** |

*Note:* At each threshold shown, we repeated the full categorization procedure, replacing ≥7/10 with the threshold to define severe symptoms; all other rules remained unchanged. Agreement is the proportion of patients whose label matched their primary ≥7/10 label. The ≥7/10 column is omitted because it serves as the reference; its distribution is given in Table 4.

### Threshold Sensitivity

Dominant-axis labels varied with the severity threshold applied (Table 5). Agreement with the primary ≥7/10 labels was 87.2% at ≥6/10 and 69.2% at both ≥5/10 and ≥8/10.

The relative frequencies of the two leading dominant-axis labels also changed across thresholds. At ≥5, Excitatory/Neuroinflammatory labels outnumbered CRH/Adrenergic labels 17 to 15; at ≥8, CRH/Adrenergic labels outnumbered Excitatory/Neuroinflammatory labels 19 to 6. Thus, which of these two labels was more frequent depended on the severity threshold applied.

Mixed labels were assigned to 3 patients at ≥5, 2 at ≥6, none at ≥7, and 1 at ≥8. The number of patients meeting the Low-symptom criterion increased as the threshold rose: 2 at ≥5, 6 at ≥6, 7 at ≥7, and 11 at ≥8. Because the Low-symptom rule was applied first, patients with three or fewer symptoms meeting the tested threshold were classified as Low-symptom before comparing the four axis counts.

Excluding the three patients who left more than 60% of items unanswered yielded similar results: agreement with the primary ≥7/10 labels was 69.4% at ≥5, 86.1% at ≥6, and 66.7% at ≥8, compared with 69.2%, 87.2%, and 69.2%, respectively, in the full cohort.

No patient received a Basal Ganglia–Cerebellar label at any threshold examined.

### MCAS-Overlap as an Exploratory Modifier

Twenty-two of 39 patients (56.4%) met the study’s operational criterion for MCAS-overlap: two or more of the 21 MCAS-overlap symptoms rated ≥5/10. The full item set is provided in Appendix 2.

MCAS-overlap was a broad, symptom-based exploratory construct developed for this study. It does not imply a formal diagnosis of mast cell activation syndrome or define a discrete patient subtype.

## Discussion

### Summary of Findings

This study describes 39 adults presenting to a specialty benzodiazepine taper program who reported substantial symptom burden and functional impairment. Thirty-seven were seeking to begin a taper, while two had completed tapering but remained symptomatic.

On the study-specific 233-item symptom questionnaire, patients endorsed a median of 74 symptoms, with a median of 24 rated severe. Thirty-five of 39 patients (89.7%) had at least one severe symptom in two or more of the four core questionnaire domains, demonstrating that symptom burden was generally multisystem rather than confined to a single domain. On the standardized, externally normed PROMIS-29, mean T-scores were elevated for Anxiety, Depression, Sleep Disturbance, and Fatigue and were lower for Physical Function and Ability to Participate in Social Roles and Activities, indicating reduced functioning. Pain Interference was close to the population-referenced mean.

The rule-based categorization provided an exploratory summary of where severe symptoms were concentrated within the questionnaire. CRH/Adrenergic and Excitatory/Neuroinflammatory were the most frequent dominant-axis labels, with 14 patients in each group; four were classified as Autonomic and seven as Low-symptom. No patient was classified as Basal Ganglia–Cerebellar or Mixed at the primary ≥7/10 threshold. Twenty-two patients (56.4%) also met the operational criterion for MCAS-overlap, which was treated as a cross-cutting modifier rather than a competing axis.

### Interpretation and Limitations of the Dominant-Axis Rule

The dominant-axis rule provided a simplified summary of the 233-item questionnaire. Patients with three or fewer severe symptoms across the four core axes were classified as Low-symptom. For the remaining patients, the label identified the axis containing the highest number of severe symptoms; ties were classified as Mixed. The labels therefore reflect an investigator-defined counting rule applied to the questionnaire.

The rule cannot establish that patients form discrete subtypes. It was not a clustering procedure, latent-class analysis, factor analysis, or other method designed to identify data-driven groups.

The rule was also limited by unequal axis size. The four competing axes contained different numbers of questionnaire items: CRH/Adrenergic 47, Excitatory/Neuroinflammatory 85, Autonomic 61, and Basal Ganglia–Cerebellar 19. Because the rule compared raw severe-symptom counts across axes with unequal numbers of items, it could not distinguish between greater symptom burden within a domain and denser sampling of that domain by the questionnaire. The unequal denominators therefore limit what the labels can establish.

The absence of Basal Ganglia–Cerebellar-dominant labels should also be interpreted cautiously. No patient received this label in the primary analysis or at any severity threshold examined. This does not show that motor, coordination, or tremor-related symptoms were absent from the cohort. The Basal Ganglia–Cerebellar axis had the fewest questionnaire items, and provenance review identified three omitted motor-related symptoms—muscle rigidity, muscle stiffness, and catatonia—from the administered questionnaire. These factors reduce confidence that the rule adequately represented this domain.

The threshold-sensitivity analysis provides a further reason for caution. When the severity threshold was shifted from ≥7/10 to ≥5/10 or ≥8/10, approximately one-third of patients received a different label. The relative frequency of the leading axis labels also changed across thresholds: Excitatory/Neuroinflammatory labels were more frequent at ≥5/10, whereas CRH/Adrenergic labels were more frequent at ≥8/10. Importantly, threshold changes also altered how many patients met the Low-symptom rule: 2 at ≥5/10, 6 at ≥6/10, 7 at ≥7/10, and 11 at ≥8/10.

Because the Low-symptom rule was applied before the axis comparison, part of the observed label instability reflects patients moving into or out of the Low-symptom category rather than switching from one dominant axis to another. These findings support interpreting the labels as threshold-dependent summaries of symptom burden. The present analysis cannot determine whether any stable latent subgroup structure exists independently of the counting rule.

### Biological Plausibility of the Symptom Domains

The five domains were established before patient categorization and were not derived by clustering or other structure-recovering analysis of this cohort. They were informed by literature on physiological and neurobiological processes relevant to long-term benzodiazepine exposure and withdrawal. That literature is summarized below to explain the rationale for the framework, not to present those mechanisms as findings of this study.

Benzodiazepines act as positive allosteric modulators at GABAₐ receptors. Sustained exposure is associated with compensatory neuroadaptation, including altered receptor subunit composition and trafficking [10–13] and changes in tonic inhibitory tone [14]. Experimental studies suggest that reduced GABAergic inhibition can be accompanied by increased excitability and altered N-methyl-D-aspartate (NMDA) receptor function [15,16], as well as heightened locus coeruleus noradrenergic activity, which CRH is known to promote [17–19]. Benzodiazepine withdrawal specifically has been associated with increased CRF expression and altered HPA-axis function [20–22].

These pathways cross conventional system boundaries. The locus coeruleus receives inhibitory baroreceptor input [23] and is central to arousal regulation [24]; baroreflex and vagal mechanisms contribute to cardiovascular and orthostatic regulation [25–27]. Sustained NMDA-receptor activation in microglia can drive pro-inflammatory signaling [28], and astrocytes contribute to central immune regulation [29]. CRH/CRF signaling can activate mast-cell pathways [30,31]; human mast cells can themselves synthesize and secrete CRH [32] and participate in neuroimmune signaling [33]. Separately, GABAₐ receptor subunit composition varies markedly across brain regions and neuronal populations [34], so reduced GABAergic inhibition need not affect all circuits equally. GABAergic signaling is central to basal ganglia function and movement disorders [35], GABAₐ α1 deficiency produces tremor and motor incoordination in mice [36], and benzodiazepines have been shown to alter thalamic and basal ganglia activity in patients with dystonia [37].

Because inhibitory signaling participates broadly across arousal, autonomic, sensory, motor, cognitive, and affective circuits, disturbance of inhibitory tone could plausibly produce symptoms across multiple domains simultaneously. This rationale for organizing the questionnaire into five domains is consistent with the breadth of symptoms reported in the cohort.

Table 6 summarizes the proposed processes, representative symptoms, and supporting literature for each domain. It describes processes for which there is published support and to which reported symptoms may plausibly be related. It does not assert that any individual patient’s symptoms were driven by a particular process. No physiological, biochemical, autonomic, immunological, imaging, or biomarker measurements were analyzed in this study, and the proposed mechanisms were not tested against these data. The account above therefore explains the rationale for the five-domain framework but does not establish that these mechanisms operated as described in this cohort.

**Table 6.**
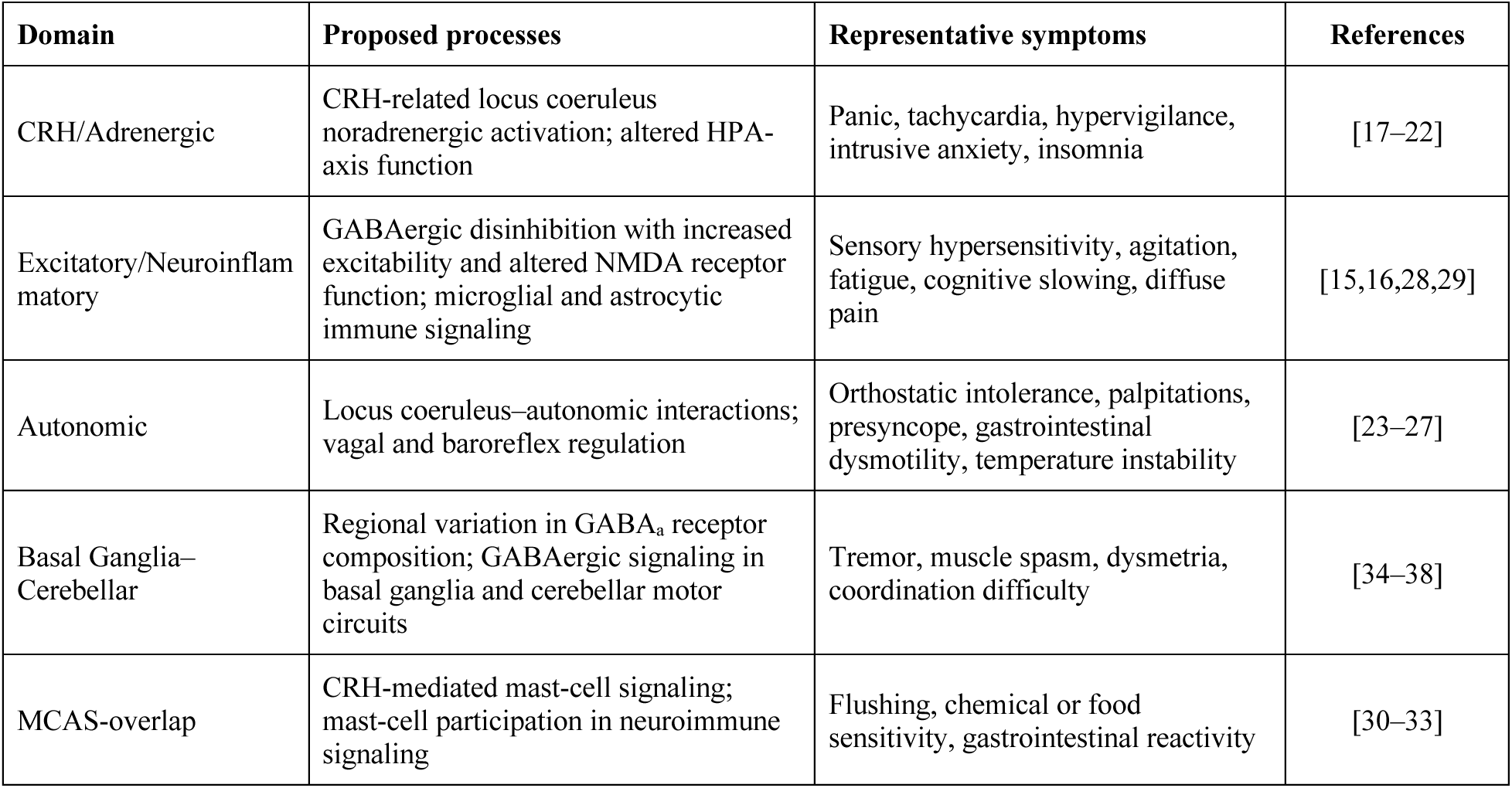
Proposed biological rationale for the five symptom domains.

| Domain | Proposed processes | Representative symptoms | References |
| --- | --- | --- | --- |
| CRH/Adrenergic | CRH-related locus coeruleus noradrenergic activation; altered HPA-axis function | Panic, tachycardia, hypervigilance, intrusive anxiety, insomnia | [17–22] |
| Excitatory/Neuroinflammatory | GABAergic disinhibition with increased excitability and altered NMDA receptor function; microglial and astrocytic immune signaling | Sensory hypersensitivity, agitation, fatigue, cognitive slowing, diffuse pain | [15,16,28,29] |
| Autonomic | Locus coeruleus–autonomic interactions; vagal and baroreflex regulation | Orthostatic intolerance, palpitations, presyncope, gastrointestinal dysmotility, temperature instability | [23–27] |
| Basal Ganglia–Cerebellar | Regional variation in GABA<sub>A</sub> receptor composition; GABAergic signaling in basal ganglia and cerebellar motor circuits | Tremor, muscle spasm, dysmetria, coordination difficulty | [34–38] |
| MCAS-overlap | CRH-mediated mast-cell signaling; mast-cell participation in neuroimmune signaling | Flushing, chemical or food sensitivity, gastrointestinal reactivity | [30–33] |

### Cohort Complexity

The cohort reported substantial psychiatric, emotional, and multisystem health complexity. All 38 patients who provided medical histories reported at least one psychiatric or emotional concern, and roughly half or more of the cohort reported health problems and concerns across neurological, endocrine–metabolic, immune, gastrointestinal, musculoskeletal, respiratory, and cardiovascular categories (Appendix 3). These findings should be interpreted in the context of prescribing patterns: benzodiazepines are commonly used for anxiety and insomnia [39]. Psychiatric history alone therefore cannot determine whether the symptoms reported here reflect pre-existing conditions, benzodiazepine withdrawal, or both.

Twenty-two of 39 patients (56.4%) met the study’s operational criterion for MCAS-overlap. Whether these symptoms reflected mast-cell or histamine-mediated processes was not assessed, and no immunological testing was performed. The frequency of this modifier is reported descriptively and does not constitute evidence of immune dysregulation.

These reported health problems and concerns were not independently verified. They are described to characterize the cohort, not to identify contributors to withdrawal severity. The study cannot determine how much of the reported symptom burden reflects benzodiazepine withdrawal versus pre-existing disease, medication effects, nutritional status, sleep disruption, stress physiology, or other contributors.

### Comparison with Prior Work

The findings are consistent with prior descriptions of benzodiazepine withdrawal as clinically heterogeneous, potentially prolonged, and often difficult to capture within a single symptom category. Ashton described benzodiazepine withdrawal as involving psychological, neurological, autonomic, gastrointestinal, sensory, and musculoskeletal symptoms, with some patients experiencing protracted symptoms after discontinuation [9]. More recent patient-survey work has similarly documented broad symptom burden and functional impairment among individuals reporting benzodiazepine-related adverse effects or withdrawal syndromes [1,2]. The present cohort similarly showed high symptom burden, multisystem symptoms, and substantial impairment on standardized functional measures.

The descriptive construct of benzodiazepine-induced neurological dysfunction, or BIND, was proposed to describe symptoms and associated functional consequences that may emerge during benzodiazepine use or tapering and continue after discontinuation [3]. The multisystem symptoms and functional impairment observed in this cohort overlap with that description.

The marked heterogeneity observed in this cohort is consistent with benzodiazepine-tapering guidance emphasizing individualized pacing and monitoring [39–41].

### Implications

The principal implication is that patients presenting for benzodiazepine taper care may have a clinically severe, multisystem presentation accompanied by substantial functional impairment. In this cohort, the clinical picture extended beyond anxiety and insomnia to include cognitive, sensory, autonomic, gastrointestinal, motor, immune-type, and pain-related symptoms. The five-domain framework provides a structured way to describe this complexity.

For patients, a structured framework may help make sense of symptoms that otherwise appear disconnected. Symptoms involving anxiety, sleep, cognition, sensation, autonomic and gastrointestinal function, movement, immune-type reactivity, and pain may occur together among patients presenting for taper care. Recognizing these patterns may support communication about complex symptom experiences. The symptom burden documented by the 233-item questionnaire and the functional impairment measured by the PROMIS-29 do not depend on dominant-axis classification.

For clinicians, the findings support comprehensive, individualized assessment. The framework may help identify areas of severe symptom burden that warrant attention, including autonomic symptoms, sensory hypersensitivity, sleep disturbance, cognitive slowing, and motor symptoms. Clinical management should remain grounded in the patient’s complete clinical presentation.

### Strengths

This study has several strengths. Patients were enrolled consecutively from a clinical practice without selection according to symptom pattern or severity. The broad 233-item symptom inventory was paired with the PROMIS-29, a standardized, population-referenced instrument, allowing detailed self-reported symptom burden to be considered alongside standardized measures of symptoms and functioning.

The analysis examined the behavior and limitations of the rule-based categorization rather than reporting its output alone. The full categorization procedure was repeated at several severity thresholds, and the implications of unequal axis sizes and the Low-symptom rule were considered explicitly. Questionnaire, medication exposure, demographic, and PROMIS data were derived from source records, with questionnaire provenance and item assignments documented in Appendix 1 and PROMIS-29 reconciliation and scoring procedures documented in the Supplement.

### Limitations

This study has several limitations related to cohort design, measurement, categorization, and construct validity.

First, the cohort was small, single-site, observational, and drawn from a self-pay specialty clinical practice. It was also demographically limited, with 73.7% of patients identifying as White and race not recorded for 15.8%. Findings may therefore not generalize to patients receiving benzodiazepine care in other clinical settings or populations. Only baseline assessments were analyzed, so the stability of symptom patterns over time could not be assessed. Without a control group, causal inference is not possible, and the study cannot determine how much of the reported symptom burden was attributable to benzodiazepine withdrawal itself versus pre-existing illness, medication effects, nutritional status, sleep disruption, stress physiology, or other contributors.

Second, symptom and functional data were entirely self-reported, and reported health problems and substance use histories were not independently verified or adjudicated. Exposure data were drawn from clinical records, but for some patients intake medication and dose were reconstructed retrospectively from multiple documented sources rather than recorded prospectively at intake.

The available records did not systematically distinguish whether patients were on stable doses, self-tapering, experiencing tolerance or interdose symptoms, or recently recovering from a dose reduction at the time of assessment, so the symptoms reported here cannot be attributed to a specific phase of benzodiazepine use or discontinuation. Together with the approximate nature of diazepam-equivalent conversion, these exposure estimates should not be interpreted as precise measurements. The broad 233-item symptom questionnaire lacks formal psychometric validation and should be interpreted as a descriptive clinical inventory rather than a validated measurement scale. PROMIS-29 domain scores were also subject to small amounts of item-level missingness, resulting in domain-specific sample sizes below 38 for Sleep Disturbance, Physical Function, and Ability to Participate.

Third, the categorization rested on investigator-defined analytic choices. The ≥7/10 severity threshold, the three-or-fewer Low-symptom criterion, and the ≥5/10 MCAS-overlap cutoff were operational conventions rather than externally validated criteria. Each questionnaire item was assigned to a single domain by the author without independent duplicate coding or blinded adjudication, although many symptoms could plausibly relate to more than one process. The dominant-axis rule also compared raw severe-symptom counts across axes containing unequal numbers of items. Approximately one-third of assignments changed when the severity threshold was moved by two points, and part of this instability reflected movement into or out of the Low-symptom category. No clustering, latent-class, factor-analytic, or other structure-recovering analysis was performed; the study therefore provides no evidence either for or against the existence of discrete patient subtypes.

Fourth, retrospective questionnaire provenance review identified four omitted source symptoms—burping, catatonia, muscle rigidity, and muscle stiffness—and one duplicated item. These were transcription artifacts rather than design decisions. Because three of the four omitted symptoms were motor-related, their absence may have reduced representation of the Basal Ganglia–Cerebellar domain and influenced the dominant-axis analysis. Consequently, the 233 administered items comprised 232 unique item labels. Because unanswered questionnaire items were treated as zero, symptom and axis counts may also have been underestimated in patients with substantial missingness, particularly when the margin between the two leading axes was small.

Finally, several study-specific constructs require caution. MCAS-overlap was based on a broad symptom set containing nonspecific features that may reflect autonomic, sensory, nutritional, dermatologic, or withdrawal-related processes rather than mast-cell activation specifically. No immunological testing was performed, and the construct should not be interpreted as a diagnosis of mast cell activation syndrome or evidence of immune dysregulation.

More broadly, no objective physiological or biological measures were analyzed, and the proposed mechanisms were not tested in this cohort. The biological literature summarized above establishes the rationale for the five-domain framework but does not demonstrate that these mechanisms operated in these patients. The findings should therefore be interpreted as descriptive and organizational rather than mechanistic, diagnostic, prognostic, or treatment-directive.

### Future Directions

Future work should determine whether stable, replicable symptom structures can be identified using clustering, latent-class, factor-analytic, network, or other multivariable approaches in substantially larger samples, and whether any such structures correspond to the domains proposed here. These analyses should account explicitly for overall symptom burden and include formal assessments of stability and reproducibility.

Future instruments would benefit from more balanced symptom coverage and formal psychometric development. The questionnaire used here was designed for clinical documentation rather than subgroup discovery and sampled the proposed domains unequally. A more systematically developed instrument could support more interpretable comparisons across symptom domains and provide a cleaner basis for future analyses of symptom structure.

Prospective longitudinal studies are needed to determine whether symptom patterns remain stable within individuals, how they change across tapering and recovery, and whether they relate to clinically meaningful outcomes such as taper tolerability or functional recovery. Testing predictive or mechanistic hypotheses will also require measures collected independently of symptom ratings, including physiological, autonomic, immunological, neurocognitive, or neuroimaging data.

Future research could also examine whether symptom patterns associated with benzodiazepine discontinuation overlap with those reported after other psychotropic medications, including selective serotonin reuptake inhibitors and antipsychotics [42–47]. Comparative studies using harmonized symptom measures and longitudinal designs could determine whether these syndromes share symptom structures, clinical trajectories, or biological correlates.

## Conclusion

Severe, multisystem symptom burden and substantial functional impairment were prominent in this cohort of adults seeking specialty benzodiazepine taper care. Anxiety and insomnia alone did not capture the breadth of their clinical presentations.

The five-domain framework offers a structured language for describing this complexity. Dominant-axis labels summarize where severe symptoms are concentrated; they do not define discrete patient subtypes.

The breadth of symptoms and functional impairment observed in this cohort underscores the importance of assessing both in patients presenting for benzodiazepine taper care.

## Supporting information

Supplement 1. PROMIS-29 Scoring Procedure and Data Quality

## List of Abbreviations

BG/Cer: Basal Ganglia–Cerebellar
BIND: Benzodiazepine-induced neurological dysfunction
CRF: Corticotropin-releasing factor
CRH: Corticotropin-releasing hormone
ENI: Excitatory–Neuroinflammatory
GABAₐ: Gamma-aminobutyric acid type A
HPA: Hypothalamic–pituitary–adrenal
IQR: Interquartile range
MCAS: Mast Cell Activation Syndrome
NMDA: N-methyl-D-aspartate
PDMP: Prescription drug monitoring program
PROMIS-29: Patient-Reported Outcomes Measurement Information System Profile v2.0
SD: Standard deviation

## Acknowledgements

The author acknowledges the patients whose clinical experiences and questionnaire responses formed the basis of this analysis. The author sincerely thanks Khristine Anne Garcia for her tireless assistance and unwavering support. Her dedicated efforts in patient care and data organization were essential to this study. The author also thanks the Benzodiazepine Information Coalition (Benzoinfo.com) for making available the comprehensive symptom list from which the 233-item withdrawal questionnaire was derived.

## Funding

Independent clinical research; no external funding.

## Conflicts of Interest

The author is the founder and clinical director of Brain Body Medical (The Benzo Taper Doctor), the practice from which this cohort was drawn.

## Ethics Statement

This study was determined to be exempt from IRB review by the Institutional Review Board of the Biomedical Research Alliance of New York (BRANY) on 3 October 2025, under 45 CFR 46.104(d) category (4)(ii) (BRANY IRB File #25-12-541-2328; determination #262681). A full waiver of HIPAA authorization was granted under 45 CFR 164.512(i)(2)(ii). All analyses were performed on de-identified data files.

## Data Availability

De-identified data underlying this manuscript are available from the corresponding author on reasonable request.

## Appendices

## Appendix 1. Questionnaire provenance and administered 233-item symptom questionnaire Derivation of the 233-item questionnaire from the source vocabulary

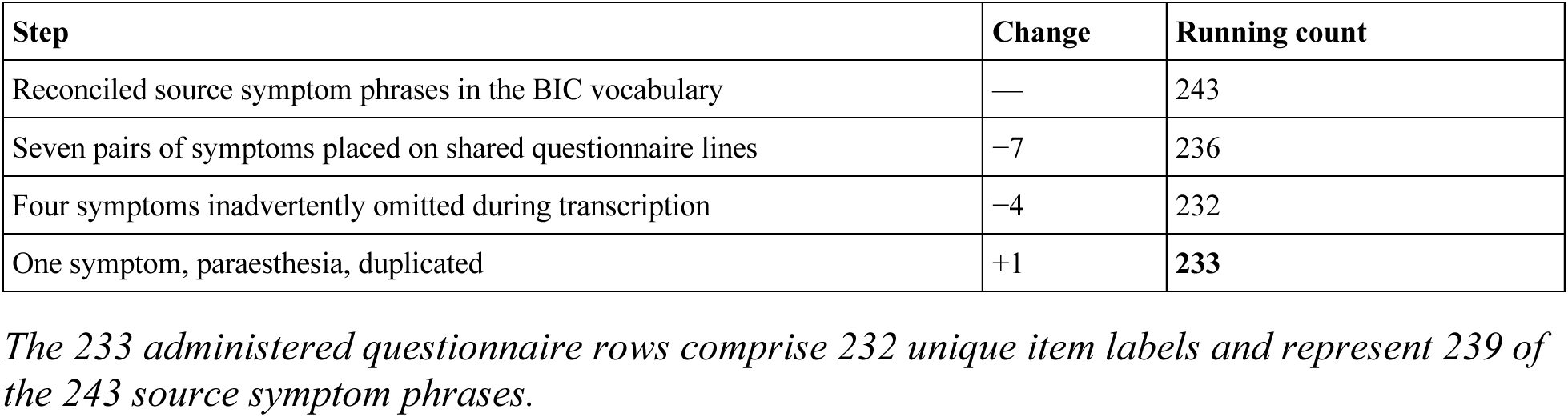

Appendix 1. Questionnaire provenance and administered 233-item symptom questionnaire Derivation of the 233-item questionnaire from the source vocabulary.
| Step | Change | Running count |
| --- | --- | --- |
| Reconciled source symptom phrases in the BIC vocabulary | — | 243 |
| Seven pairs of symptoms placed on shared questionnaire lines | −7 | 236 |
| Four symptoms inadvertently omitted during transcription | −4 | 232 |
| One symptom, paraesthesia, duplicated | +1 | **233** |

*The 233 administered questionnaire rows comprise 232 unique item labels and represent 239 of the 243 source symptom phrases.*

### The administered 233-item questionnaire

All 233 rows are listed below in the order administered, with the domain to which each item was assigned for the rule-based count. Patients rated every item from 0 to 10. Ordering is broadly alphabetical but not strictly so; the duplicated paraesthesia row appears at position 197 as noted below.

| Axis | Items |
| --- | --- |
| Excitatory/Neuroinflammatory | 85 |
| Autonomic | 61 |
| CRH/Adrenergic | 47 |
| Basal Ganglia–Cerebellar | 19 |
| MCAS-overlap (cross-cutting modifier, not competing for the dominant-axis label) | 21 |
| Total | 233 |

| # | Questionnaire item (rated 0–10) | Assigned domain |
| --- | --- | --- |
| 1 | Abdomen Cramping and Distention | Autonomic |
| 2 | Achiness | Excitatory/Neuroinflammatory |
| 3 | Aching jaw | Excitatory/Neuroinflammatory |
| 4 | Aching joints | Excitatory/Neuroinflammatory |
| 5 | Aching muscles | Excitatory/Neuroinflammatory |
| 6 | Acute hyperexcited state | CRH/Adrenergic |
| 7 | Aggressive behaviour | CRH/Adrenergic |
| 8 | Agitation | CRH/Adrenergic |
| 9 | Agitated sight | Excitatory/Neuroinflammatory |
| 10 | Agoraphobia | CRH/Adrenergic |
| 11 | Air Hunger | Autonomic |
| 12 | Akathisia | Basal Ganglia–Cerebellar |
| 13 | Allergic reactions | MCAS-overlap |
| 14 | Allergic reactions to/Intolerance of foods previously tolerated | MCAS-overlap |
| 15 | Allergy and nasal symptoms exacerbated | MCAS-overlap |
| 16 | Anger | CRH/Adrenergic |
| 17 | Anhedonia | Excitatory/Neuroinflammatory |
| 18 | Anorgasmia | Autonomic |
| 19 | Anxiety | CRH/Adrenergic |
| 20 | Apathy | Excitatory/Neuroinflammatory |
| 21 | Appetite - either loss of, or voracious appetite with constant desire to eat | CRH/Adrenergic |
| 22 | Arms and legs feel detached from body | CRH/Adrenergic |
| 23 | Ataxia | Basal Ganglia–Cerebellar |
| 24 | Band around head | CRH/Adrenergic |
| 25 | Bladder Incontinence | Autonomic |
| 26 | Bleeding between menstrual cycles | CRH/Adrenergic |
| 27 | Bleeding from the nose | MCAS-overlap |
| 28 | Blepharospasm | Basal Ganglia–Cerebellar |
| 29 | Blood Pressure Issues (High or Low) | Autonomic |
| 30 | Blurred Vision | Autonomic |
| 31 | Body Temperature Fluctuations | MCAS-overlap |
| 32 | Brain Fog | Excitatory/Neuroinflammatory |
| 33 | Body odor | Autonomic |
| 34 | Bradycardia | Autonomic |
| 35 | Brain moving within the skull sensation | Excitatory/Neuroinflammatory |
| 36 | Breasts (heavy, over-sensitive, swollen, enlarged, painful) | Excitatory/Neuroinflammatory |
| 37 | Breathing difficulties | Autonomic |
| 38 | Breathlessness | Autonomic |
| 39 | Bruxism | Basal Ganglia–Cerebellar |
| 40 | Buildings appear to be leaning | Excitatory/Neuroinflammatory |
| 41 | Burning along the spine | Excitatory/Neuroinflammatory |
| 42 | Burning Sensations on Skin | Excitatory/Neuroinflammatory |
| 43 | Buzzing, burning, tingling limbs | Excitatory/Neuroinflammatory |
| 44 | Changes in perception (faces distorting and inanimate objects moving) | CRH/Adrenergic |
| 45 | Changes in skin color, tone, texture | MCAS-overlap |
| 46 | Chemical Sensitivities | MCAS-overlap |
| 47 | Chest tightness | Autonomic |
| 48 | Chills | Autonomic |
| 49 | Choking | Autonomic |
| 50 | Claustrophobia | CRH/Adrenergic |
| 51 | Clumsiness | Basal Ganglia–Cerebellar |
| 52 | Cognitive impairment | Excitatory/Neuroinflammatory |
| 53 | Confusion | Excitatory/Neuroinflammatory |
| 54 | Constipation | Autonomic |
| 55 | Cracked and sore lips | Autonomic |
| 56 | Cramping in stomach | Autonomic |
| 57 | Cramping of the muscles | Excitatory/Neuroinflammatory |
| 58 | Craving for sweets and sugary foods | CRH/Adrenergic |
| 59 | Crying Spells | Excitatory/Neuroinflammatory |
| 60 | Cuts and abrasions taking weeks to heal | MCAS-overlap |
| 61 | Dark circles under eyes | MCAS-overlap |
| 62 | Decaying teeth and gums | MCAS-overlap |
| 63 | Dehydration | Autonomic |
| 64 | Delirium | Excitatory/Neuroinflammatory |
| 65 | Demented and murderous thoughts | Excitatory/Neuroinflammatory |
| 66 | Dental Pain | Excitatory/Neuroinflammatory |
| 67 | Depersonalization | Excitatory/Neuroinflammatory |
| 68 | Depression | Excitatory/Neuroinflammatory |
| 69 | Diarrhea | Autonomic |
| 70 | Difficulty swallowing | Autonomic |
| 71 | Disorientation | Excitatory/Neuroinflammatory |
| 72 | Dizziness | Autonomic |
| 73 | Double Vision | Excitatory/Neuroinflammatory |
| 74 | Dry Retching | Autonomic |
| 75 | Dry throat, sore tongue, and thrush | Autonomic |
| 76 | Dyspepsia | Autonomic |
| 77 | Dysphagia (difficulty eating or swallowing) | Autonomic |
| 78 | Dysphoria (inability to feel pleasure or happiness) | Excitatory/Neuroinflammatory |
| 79 | Dyspnea (breathing difficulty) | Autonomic |
| 80 | Dystonia | Basal Ganglia–Cerebellar |
| 81 | Earache and sinus problems | MCAS-overlap |
| 82 | Edema | Autonomic |
| 83 | Electronic Zaps | Excitatory/Neuroinflammatory |
| 84 | Emotional Blunting | Excitatory/Neuroinflammatory |
| 85 | Epileptic Fits | Excitatory/Neuroinflammatory |
| 86 | Esophageal Spasms | Autonomic |
| 87 | Esophagitis | Autonomic |
| 88 | Excitability | CRH/Adrenergic |
| 89 | Exhaustion | CRH/Adrenergic |
| 90 | Extreme cold | Autonomic |
| 91 | Extreme thirst | Autonomic |
| 92 | Eyes - sore, tired, seeing floaters, spots, itchy, red | Excitatory/Neuroinflammatory |
| 93 | Eye Dryness | Autonomic |
| 94 | Facial Numbness | Excitatory/Neuroinflammatory |
| 95 | Fear (terror) | CRH/Adrenergic |
| 96 | Feeling of extreme heat or cold | MCAS-overlap |
| 97 | Feeling of impending doom | CRH/Adrenergic |
| 98 | Feelings of shaking inside and out | CRH/Adrenergic |
| 99 | Feeling of shame | Excitatory/Neuroinflammatory |
| 100 | Feelings of unreality | CRH/Adrenergic |
| 101 | Feelings of worms under scalp | MCAS-overlap |
| 102 | Female ejaculation or incontinence during orgasm | Autonomic |
| 103 | Fingernail problems - median nail dystrophy (line/ridge down the centre), ridges | MCAS-overlap |
| 104 | Flashbacks | Excitatory/Neuroinflammatory |
| 105 | Fluctuations in blood pressure | Autonomic |
| 106 | Flu-like symptoms (“Benzo flu”) | Excitatory/Neuroinflammatory |
| 107 | Flushing | MCAS-overlap |
| 108 | Formication | MCAS-overlap |
| 109 | Fuzzy feeling in head | Excitatory/Neuroinflammatory |
| 110 | Gait Abnormality | Basal Ganglia–Cerebellar |
| 111 | Gastritis | MCAS-overlap |
| 112 | Glassy eyes | Autonomic |
| 113 | Goosebumps - very visible to the eye | MCAS-overlap |
| 114 | Grinding teeth | Basal Ganglia–Cerebellar |
| 115 | Hair Changes (loss, thinning, dullness) | MCAS-overlap |
| 116 | Hallucinations | Excitatory/Neuroinflammatory |
| 117 | Headaches | Excitatory/Neuroinflammatory |
| 118 | Heartburn | Autonomic |
| 119 | Heavy arms and legs | Autonomic |
| 120 | Hemorrhoids | Autonomic |
| 121 | Holding onto the walls in the shower | Excitatory/Neuroinflammatory |
| 122 | Hypersensitive Senses | Excitatory/Neuroinflammatory |
| 123 | Heart Palpitations | Autonomic |
| 124 | Hormonal Imbalances | CRH/Adrenergic |
| 125 | Hot Flashes | Autonomic |
| 126 | Hurts to wear clothes, a car seatbelt | Excitatory/Neuroinflammatory |
| 127 | Hypersensitive (unable to watch the news, see films, read the newspaper) | Excitatory/Neuroinflammatory |
| 128 | Hyperactivity | CRH/Adrenergic |
| 129 | Hyperacusis (profound hearing sensitivity, hurts to talk on the phone, music, etc.) | Excitatory/Neuroinflammatory |
| 130 | Hypersomia (sensitive sense of smell) | Excitatory/Neuroinflammatory |
| 131 | Hyperreflexia ('jumpiness') Hypersensitivity to being touched | Basal Ganglia–Cerebellar |
| 132 | Hypersensitivity to light, sound, and other stimuli | Excitatory/Neuroinflammatory |
| 133 | Hypersensitivity to stress | CRH/Adrenergic |
| 134 | Hypertension (elevated blood pressure) | Autonomic |
| 135 | Hyperventilation (over breathing) | Autonomic |
| 136 | Hypnologic (hallucinations, sleepwalking) | CRH/Adrenergic |
| 137 | Hysterical and inappropriate laughter | Excitatory/Neuroinflammatory |
| 138 | IBS - irritable bowel syndrome | Autonomic |
| 139 | Impacted bowels | Autonomic |
| 140 | Impairment of motor coordination Impotence | Basal Ganglia–Cerebellar |
| 141 | Inability to comprehend the simplest things | Excitatory/Neuroinflammatory |
| 142 | Inability to concentrate | Excitatory/Neuroinflammatory |
| 143 | Inability to cope with a lot of information Inability to draw a satisfying breath | Excitatory/Neuroinflammatory |
| 144 | Inability to make a decision | CRH/Adrenergic |
| 145 | Inability to read or write | Excitatory/Neuroinflammatory |
| 146 | Incontinence, frequent or urgent need to urinate, unable to hold or pass urine | Autonomic |
| 147 | Increased saliva | Autonomic |
| 148 | Indigestion | Autonomic |
| 149 | Insomnia | CRH/Adrenergic |
| 150 | Intense fuzzy feeling in the head | Excitatory/Neuroinflammatory |
| 151 | Intense jaw pain | Excitatory/Neuroinflammatory |
| 152 | Intrusive Thoughts | CRH/Adrenergic |
| 153 | Iris in eyes changes color | Excitatory/Neuroinflammatory |
| 154 | Irrational rage | CRH/Adrenergic |
| 155 | Irrationality (in general) Irritability | CRH/Adrenergic |
| 156 | Jaundice | Excitatory/Neuroinflammatory |
| 157 | Jaw, tooth, neck and shoulder aches and pains | Excitatory/Neuroinflammatory |
| 158 | Jaw tension, clenching, teeth grinding | Basal Ganglia–Cerebellar |
| 159 | “Jelly Legs” - weakness in the legs | Autonomic |
| 160 | Jerks in arms, legs fingers | Basal Ganglia–Cerebellar |
| 161 | Joint aches and pains | Excitatory/Neuroinflammatory |
| 162 | Joylessness | Excitatory/Neuroinflammatory |
| 163 | Jumpiness | CRH/Adrenergic |
| 164 | Lack of concentration Lack of coordination | Excitatory/Neuroinflammatory |
| 165 | Left/Right-sided symptoms | Excitatory/Neuroinflammatory |
| 166 | Legs arms and head very heavy Leukonychia (whitening of nails) | Autonomic |
| 167 | Libidinal changes - variances in sex drive | Autonomic |
| 168 | Loss of balance | Autonomic |
| 169 | Loss of concentration | Excitatory/Neuroinflammatory |
| 170 | Loss of confidence | CRH/Adrenergic |
| 171 | Loss of control of movement | Basal Ganglia–Cerebellar |
| 172 | Loss of interest in people and/or things Loss of memory (short and/or long-term) | Excitatory/Neuroinflammatory |
| 173 | Loss of self respect | Excitatory/Neuroinflammatory |
| 174 | Loss of taste or metallic taste (or other strange tastes) | Excitatory/Neuroinflammatory |
| 175 | Loss or changes in appetite | CRH/Adrenergic |
| 176 | Lung pain | Excitatory/Neuroinflammatory |
| 177 | Memory Problems | Excitatory/Neuroinflammatory |
| 178 | Menstrual Problems | CRH/Adrenergic |
| 179 | Metallic Taste | Excitatory/Neuroinflammatory |
| 180 | Mood swings | Excitatory/Neuroinflammatory |
| 181 | Muscle aches | Excitatory/Neuroinflammatory |
| 182 | Muscle spasms | Basal Ganglia–Cerebellar |
| 183 | Muscle weakness | Autonomic |
| 184 | Nausea | Autonomic |
| 185 | Nervousness | CRH/Adrenergic |
| 186 | Nightmares | CRH/Adrenergic |
| 187 | Night sweats | CRH/Adrenergic |
| 188 | Numbness/Tingling | Excitatory/Neuroinflammatory |
| 189 | Obsessions and obsessive behavior, OCD | CRH/Adrenergic |
| 190 | Orgasm changes | Autonomic |
| 191 | Outbursts of aggression and rage | CRH/Adrenergic |
| 192 | Pain | Excitatory/Neuroinflammatory |
| 193 | Painful scalp | Excitatory/Neuroinflammatory |
| 194 | Panic Attacks | CRH/Adrenergic |
| 195 | Paraesthesia | Excitatory/Neuroinflammatory |
| 196 | Paranoia | Excitatory/Neuroinflammatory |
| 197 | Paraesthesia | Excitatory/Neuroinflammatory |
| 198 | Parosmia | Excitatory/Neuroinflammatory |
| 199 | Perception Distortion | Excitatory/Neuroinflammatory |
| 200 | Passing gas continually | Autonomic |
| 201 | Phobias | CRH/Adrenergic |
| 202 | Post Traumatic Stress symptoms | CRH/Adrenergic |
| 203 | Postural orthostatic tachycardia syndrome (POTS) | Autonomic |
| 204 | Premature menopause | CRH/Adrenergic |
| 205 | Pseudobulbar / PBA (uncontrollable crying or laugh) | Excitatory/Neuroinflammatory |
| 206 | Psychotic symptoms | Excitatory/Neuroinflammatory |
| 207 | Racing Thoughts | CRH/Adrenergic |
| 208 | Rapid blinking of the eyes | Basal Ganglia–Cerebellar |
| 209 | Rapid weight loss | CRH/Adrenergic |
| 210 | Rashes on skin | MCAS-overlap |
| 211 | Reading/Comprehension Problems | Excitatory/Neuroinflammatory |
| 212 | Repetative Thoughts | CRH/Adrenergic |
| 213 | Restless Legs | Basal Ganglia–Cerebellar |
| 214 | Scalp Pain | Excitatory/Neuroinflammatory |
| 215 | Shivering | Autonomic |
| 216 | Sinusitis | MCAS-overlap |
| 217 | Speech difficulties | Excitatory/Neuroinflammatory |
| 218 | Swallowing difficulties | Autonomic |
| 219 | Tachycardia | Autonomic |
| 220 | Teeth Pain | Excitatory/Neuroinflammatory |
| 221 | Thyroid disturbances | CRH/Adrenergic |
| 222 | Ticking | Basal Ganglia–Cerebellar |
| 223 | Tinnitus | Excitatory/Neuroinflammatory |
| 224 | Toxic naps | CRH/Adrenergic |
| 225 | Tremor | Basal Ganglia–Cerebellar |
| 226 | Twitching | Basal Ganglia–Cerebellar |
| 227 | Urinary Problems | Autonomic |
| 228 | Vertigo | Excitatory/Neuroinflammatory |
| 229 | Visual disturbances - waves, sparks, flashes of light, seeing spots, floaters, blurry | Excitatory/Neuroinflammatory |
| 230 | Vomiting | Autonomic |
| 231 | Weakness | Autonomic |
| 232 | Weight Gain or Loss | CRH/Adrenergic |
| 233 | Zaps | Excitatory/Neuroinflammatory |

*Note:* Patients rated each symptom for severity on a 0–10 numeric scale. This questionnaire was administered clinically; it is not a psychometrically validated research instrument. Four source symptoms — burping, catatonia, muscle rigidity, and muscle stiffness — were inadvertently omitted when the source vocabulary was transcribed into the 233-item clinical questionnaire. Three of the four are motor-related, which may have modestly reduced motor-domain representation in the rule-based analysis. A further item (paraesthesia) is duplicated within the 233 columns, so the 233 administered rows comprise 232 unique item labels and represent 239 of the 243 source symptom phrases.

## Appendix 2. MCAS-overlap item set and operational definition

| Tier | Number of symptoms | Patients meeting tier criterion (N = 39) |
| --- | --- | --- |
| Tier 1 – Hallmark | 5 | 11 (28.2%) |
| Tier 2 – Supportive | 12 | 13 (33.3%) |
| Tier 3 – Peripheral | 4 | 10 (25.6%) |

*Note:* Percentages reflect patients with $\geq 2$ symptoms in a tier rated $\geq 5/10$ in severity. Tiers are reported separately; the overall MCAS-overlap rate (22/39, 56.4%) pools symptoms across all three tiers. Tiers are not mutually exclusive; a single patient may meet criteria for more than one tier. MCAS-overlap refers to a symptom-based modifier and does not represent a formal MCAS diagnosis.

### Symptom lists (from the 233-item symptom questionnaire)

- **Tier 1 – Hallmark (5):** Allergic reactions; Allergic reactions to/Intolerance of foods previously tolerated; Allergy and nasal symptoms exacerbated; Flushing; Sinusitis.
- **Tier 2 – Supportive (12):** Rashes on skin; Chemical Sensitivities; Gastritis; Bleeding from the nose; Changes in skin color, tone, texture; Dark circles under eyes; Cuts and abrasions taking weeks to heal; Decaying teeth and gums; Earache and sinus problems; Goosebumps - very visible to the eye; Hair Changes (loss, thinning, dullness); Fingernail problems - median nail dystrophy (line/ridge down the center), ridges.
- **Tier 3 – Peripheral (4):** Body Temperature Fluctuations; Feeling of extreme heat or cold; Feelings of worms under scalp; Formication.

## Appendix 3. Self-reported health problems and concerns by body system

| **Body system** | **N (out of 38)** | **Percent (%)** |
| --- | --- | --- |
| Psychiatric / Mental Health | 38 | 100.0 |
| Neurological | 27 | 71.1 |
| Endocrine / Metabolic | 24 | 63.2 |
| Immune / Autoimmune | 22 | 57.9 |
| Respiratory | 20 | 52.6 |
| Gastrointestinal | 22 | 57.9 |
| Cardiovascular | 19 | 50.0 |
| Musculoskeletal | 21 | 55.3 |

## Appendix 4. Substance-use history at intake

| Substance | Current n (%) | Past n (%) | Never n (%) |
| --- | --- | --- | --- |
| Benzodiazepines | 34 (94.4%) | 2 (5.6%) | 0 (0.0%) |
| Alcohol | 9 (25.0%) | 12 (33.3%) | 15 (41.7%) |
| Tobacco/Nicotine | 8 (22.2%) | 14 (38.9%) | 14 (38.9%) |
| Cannabis | 8 (22.2%) | 12 (33.3%) | 16 (44.4%) |
| Opioids | 4 (11.1%) | 3 (8.3%) | 29 (80.6%) |
| Stimulants | 2 (5.6%) | 5 (13.9%) | 29 (80.6%) |
| Food/sugar | 11 (30.6%) | 14 (38.9%) | 11 (30.6%) |

*Note:* Any reported current use—regardless of frequency or quantity, including occasional use—was coded as “current.” Data were available for 36 of 39 patients; three (BenzoQ36, BenzoQ30, BenzoQ34) did not provide this information.

## Appendix 5. Patient-level dominant-axis tallies (N = 39)

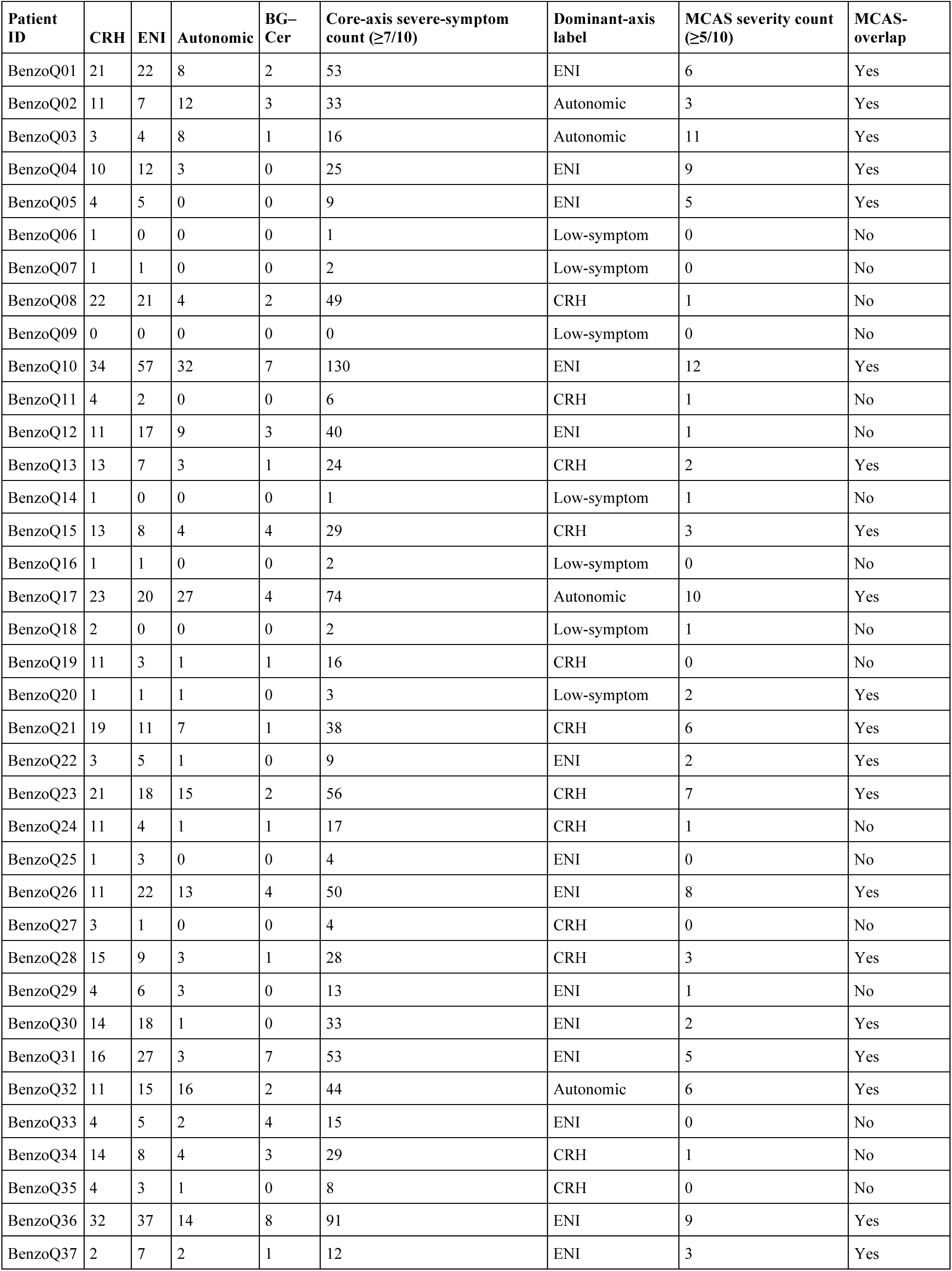
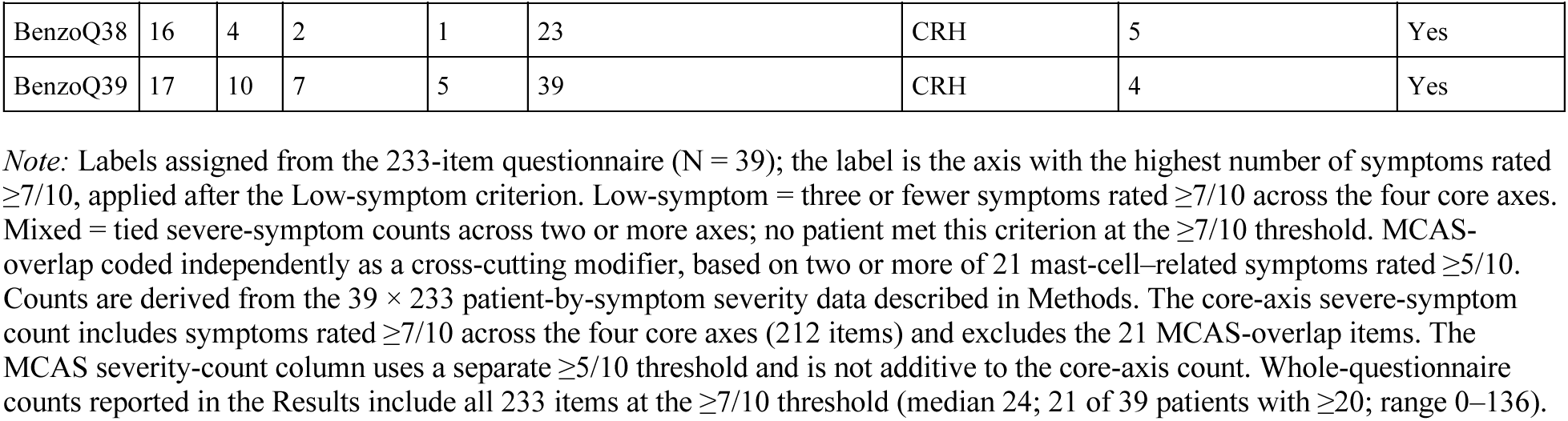

| Patient ID | CRH | ENI | Autonomic | BG–Cer | Core-axis severe-symptom count ($\geq 7/10$) | Dominant-axis label | MCAS severity count ($\geq 5/10$) | MCAS-overlap |
| --- | --- | --- | --- | --- | --- | --- | --- | --- |
| BenzoQ01 | 21 | 22 | 8 | 2 | 53 | ENI | 6 | Yes |
| BenzoQ02 | 11 | 7 | 12 | 3 | 33 | Autonomic | 3 | Yes |
| BenzoQ03 | 3 | 4 | 8 | 1 | 16 | Autonomic | 11 | Yes |
| BenzoQ04 | 10 | 12 | 3 | 0 | 25 | ENI | 9 | Yes |
| BenzoQ05 | 4 | 5 | 0 | 0 | 9 | ENI | 5 | Yes |
| BenzoQ06 | 1 | 0 | 0 | 0 | 1 | Low-symptom | 0 | No |
| BenzoQ07 | 1 | 1 | 0 | 0 | 2 | Low-symptom | 0 | No |
| BenzoQ08 | 22 | 21 | 4 | 2 | 49 | CRH | 1 | No |
| BenzoQ09 | 0 | 0 | 0 | 0 | 0 | Low-symptom | 0 | No |
| BenzoQ10 | 34 | 57 | 32 | 7 | 130 | ENI | 12 | Yes |
| BenzoQ11 | 4 | 2 | 0 | 0 | 6 | CRH | 1 | No |
| BenzoQ12 | 11 | 17 | 9 | 3 | 40 | ENI | 1 | No |
| BenzoQ13 | 13 | 7 | 3 | 1 | 24 | CRH | 2 | Yes |
| BenzoQ14 | 1 | 0 | 0 | 0 | 1 | Low-symptom | 1 | No |
| BenzoQ15 | 13 | 8 | 4 | 4 | 29 | CRH | 3 | Yes |
| BenzoQ16 | 1 | 1 | 0 | 0 | 2 | Low-symptom | 0 | No |
| BenzoQ17 | 23 | 20 | 27 | 4 | 74 | Autonomic | 10 | Yes |
| BenzoQ18 | 2 | 0 | 0 | 0 | 2 | Low-symptom | 1 | No |
| BenzoQ19 | 11 | 3 | 1 | 1 | 16 | CRH | 0 | No |
| BenzoQ20 | 1 | 1 | 1 | 0 | 3 | Low-symptom | 2 | Yes |
| BenzoQ21 | 19 | 11 | 7 | 1 | 38 | CRH | 6 | Yes |
| BenzoQ22 | 3 | 5 | 1 | 0 | 9 | ENI | 2 | Yes |
| BenzoQ23 | 21 | 18 | 15 | 2 | 56 | CRH | 7 | Yes |
| BenzoQ24 | 11 | 4 | 1 | 1 | 17 | CRH | 1 | No |
| BenzoQ25 | 1 | 3 | 0 | 0 | 4 | ENI | 0 | No |
| BenzoQ26 | 11 | 22 | 13 | 4 | 50 | ENI | 8 | Yes |
| BenzoQ27 | 3 | 1 | 0 | 0 | 4 | CRH | 0 | No |
| BenzoQ28 | 15 | 9 | 3 | 1 | 28 | CRH | 3 | Yes |
| BenzoQ29 | 4 | 6 | 3 | 0 | 13 | ENI | 1 | No |
| BenzoQ30 | 14 | 18 | 1 | 0 | 33 | ENI | 2 | Yes |
| BenzoQ31 | 16 | 27 | 3 | 7 | 53 | ENI | 5 | Yes |
| BenzoQ32 | 11 | 15 | 16 | 2 | 44 | Autonomic | 6 | Yes |
| BenzoQ33 | 4 | 5 | 2 | 4 | 15 | ENI | 0 | No |
| BenzoQ34 | 14 | 8 | 4 | 3 | 29 | CRH | 1 | No |
| BenzoQ35 | 4 | 3 | 1 | 0 | 8 | CRH | 0 | No |
| BenzoQ36 | 32 | 37 | 14 | 8 | 91 | ENI | 9 | Yes |
| BenzoQ37 | 2 | 7 | 2 | 1 | 12 | ENI | 3 | Yes |

| Patient ID | CRH | ENI | Autonomic | BG-Cer | Core-axis severe-symptom count ($\geq 7/10$) | Dominant-axis label | MCAS severity count ($\geq 5/10$) | MCAS-overlap |
| --- | --- | --- | --- | --- | --- | --- | --- | --- |
| BenzoQ38 | 16 | 4 | 2 | 1 | 23 | CRH | 5 | Yes |
| BenzoQ39 | 17 | 10 | 7 | 5 | 39 | CRH | 4 | Yes |

*Note:* Labels assigned from the 233-item questionnaire (N = 39); the label is the axis with the highest number of symptoms rated $\geq 7/10$ , applied after the Low-symptom criterion. Low-symptom = three or fewer symptoms rated $\geq 7/10$ across the four core axes. Mixed = tied severe-symptom counts across two or more axes; no patient met this criterion at the $\geq 7/10$ threshold. MCAS-overlap coded independently as a cross-cutting modifier, based on two or more of 21 mast-cell-related symptoms rated $\geq 5/10$ . Counts are derived from the $39 \times 233$ patient-by-symptom severity data described in Methods. The core-axis severe-symptom count includes symptoms rated $\geq 7/10$ across the four core axes (212 items) and excludes the 21 MCAS-overlap items. The MCAS severity-count column uses a separate $\geq 5/10$ threshold and is not additive to the core-axis count. Whole-questionnaire counts reported in the Results include all 233 items at the $\geq 7/10$ threshold (median 24; 21 of 39 patients with $\geq 20$ ; range 0–136).

## Appendix 6. Data completeness at intake

| Instrument | Completed (n) | Missing (n) | Notes |
| --- | --- | --- | --- |
| 233-item symptom questionnaire | 39 (100%) | 0 | Completed by all 39 patients <sup>a</sup> |
| PROMIS-29 | 38 (97.4%) | 1 | Missing for BenzoQ29 <sup>b, c</sup> |
| Substance use history | 36 (92.3%) | 3 | Missing for BenzoQ30, BenzoQ34, BenzoQ36 <sup>c</sup> |
| Medical history | 38 (97.4%) | 1 | Missing for BenzoQ35 <sup>c</sup> |
| Dominant-axis label assignment | 39 (100%) | 0 | Derived from the 233-item questionnaire |

*Footnotes:* <sup>a</sup> The median proportion of unanswered items was 1.3% (see Results). <sup>b</sup> Patient identifiers are anonymized as BenzoQ codes. <sup>c</sup> Missing data reflect non-completion of forms.

## Notes

### Author Declarations

The Institutional Review Board of the Biomedical Research Alliance of New York (BRANY) waived ethical approval for this work.

### Summary of Updates

Interpretation of the rule-based symptom categories revised to present them as exploratory clinical summaries rather than discrete biological phenotypes or guides to treatment selection; descriptive results corrected following re-review of source records, including PROMIS-29 scoring, demographics, benzodiazepine and Z drug exposure, and category counts; threshold-sensitivity analysis and expanded limitations added; title, affiliation, tables, figures, appendices, and supplemental material updated. The cohort and underlying source data are unchanged.

