## Supplement 1. PROMIS-29 Scoring Procedure and Data Quality for "Multisystem Symptom Burden and Functional Impairment in a Specialty Benzodiazepine Taper Cohort"

Supplement 1. PROMIS-29 Scoring and Data Reconciliation

### S1.1 Purpose

PROMIS-29 scores in this revision were recalculated from the source questionnaires. Differences from the prior manuscript version reflect correction of response-direction scoring, selection of the baseline administration, and source-level reconciliation of ambiguous and unanswered responses. This supplement documents the procedures used to derive the values reported in Table 3.

### S1.2 Instrument and Scoring Source

PROMIS-29 Profile v2.0 was completed by 38 of the 39 patients. Most patients used an online questionnaire template; an official PROMIS-29 PDF was used when a patient could not access the online form.

Each of the seven domains contains four items. Raw summed scores were converted to T-scores using the official conversion tables for PROMIS-29 Profile v2.0 in the HealthMeasures *Adult Profile Instruments: Scoring Manual* [7] (13 February 2020), pages 16–17. PROMIS T-scores have a population-referenced mean of 50 and standard deviation of 10. Pain Intensity was recorded separately on its original 0–10 scale.

### S1.3 Response-Direction Correction

Five PROMIS-29 items require scoring in the direction specified by the instrument:

Sleep Disturbance item 18: “My sleep was refreshing”

All four Ability to Participate in Social Roles and Activities items, each worded “I have trouble doing…”

The online questionnaire template used for most administrations printed the response values in the opposite direction from the official PROMIS form:

| **Scale** | **Official PROMIS form** | **Online template as printed** |
| --- | --- | --- |
| “My sleep was refreshing” | Not at all **(5)** → Very much **(1)** | Not at all (1) → Very much (5) |
| Ability to Participate items | Never **(5)** → Always **(1)** | Never (1) → Always (5) |

The correct scoring direction was confirmed against the official PROMIS-29 scoring documentation and a completed official PDF form from the cohort. If scored according to the values printed on the online template, the Ability to Participate scale would be inverted: a patient reporting that they “always” had trouble would receive a high ability score. All scores reported in this revision apply the response direction specified by PROMIS rather than the numeric direction printed on the online template.

### S1.4 Baseline Administration

Several source questionnaires contained more than one dated administration in repeated column blocks, reflecting serial assessments during tapering. Consistent with the cross-sectional study design, only the first, baseline administration was scored.

### S1.5 Ambiguous and Unanswered Responses

Among 1,064 baseline domain-item responses, 15 (1.4%) did not contain a single unambiguous mark.

| **Category** | **n** | **Rule applied** |
|---|---|---|
| Malformed electronic cell value | 1 | Treated as a response; value recovered |
| Two adjacent response options marked | 8 | Resolved to the more severe option |
| Two nonadjacent response options marked | 1 | Resolved to the more severe option |
| No response option marked | 5 | Left unanswered; domain not scored for that patient |

A sensitivity analysis compared three approaches to the nine ambiguous responses: excluding them, resolving them to the more severe option, and resolving them to the less severe option. Domain means differed by no more than 0.2 T-score points across these approaches.

| **Domain** | **Exclude ambiguous** | **More severe** | **Less severe** |
|---|---|---|---|
| Anxiety | 36 / 68.8 | 38 / 68.8 | 38 / 68.7 |
| Depression | 37 / 62.5 | 38 / 62.3 | 38 / 62.3 |
| Sleep Disturbance | 34 / 60.9 | 36 / 61.0 | 36 / 60.9 |
| Fatigue | 38 / 60.7 | 38 / 60.7 | 38 / 60.7 |
| Pain Interference | 38 / 51.5 | 38 / 51.5 | 38 / 51.5 |
| Physical Function | 36 / 45.5 | 37 / 45.5 | 37 / 45.4 |

| Domain | Exclude ambiguous | More severe | Less severe |
|---|---|---|---|
| Ability to Participate | 36 / 40.6 | 37 / 40.7 | 37 / 40.8 |

*n / mean T-score. The “More severe” column corresponds to the values reported in Table 3.*

The conversion tables require complete responses for all four items within a domain. A domain containing an unanswered item was therefore not scored for that patient.

| Patient | Domain | Unanswered items |
|---|---|---|
| BenzoQ07 | Physical Function | 1 |
| BenzoQ20 | Sleep Disturbance | 2 |
| BenzoQ23 | Ability to Participate in Social Roles and Activities | 1 |
| BenzoQ36 | Sleep Disturbance | 1 |

These responses were rechecked against the source questionnaires and confirmed as unanswered at baseline. Responses recorded at later administrations were not substituted for missing baseline responses.

S1.6 Scoring Documentation

Scoring was performed using a documented script that applied the procedures described above to the source questionnaires. The script, transcribed conversion tables, and patient-level scores are maintained as versioned files and can be made available for independent verification, subject to the participant-privacy constraints described in the Data Availability statement.
